# Investigation of the causative pathogen in the 2023 conjunctivitis outbreak of Nepal using unbiased metagenomic next generation sequencing

**DOI:** 10.1101/2024.04.16.24305920

**Authors:** Eliya Shrestha, Nishan Katuwal, Ranju Kharel Sitaula, Harimaya Gurung, Aastha Shrestha, Pratap Karki, Rajeev Shrestha

**Author notes:** **Corresponding Author:** Prof. Dr. Rajeev Shrestha. equal contribution (Eliya Shrestha, Nishan Katuwal).

## Abstract

In mid-2023, Nepal experienced a significant outbreak of conjunctivitis, affecting over 60% of outpatients in eye hospitals and prompting school closures. The outbreak, peaking in August, predominantly impacted children and individuals with compromised immunity. Clinical manifestations included sudden-onset redness, foreign body sensation, watery discharge, and occasional lid swelling. A majority of cases exhibited acute hemorrhagic conjunctivitis, with management involving ocular lubricants, personal hygiene, and topical antibiotics. This study details the genomic epidemiology and clinical characteristics of conjunctivitis cases during the outbreak. To understand the causative agents, conjunctival swabs from patients were evaluated using unbiased metagenomic next-generation sequencing (mNGS) in Illumina iSeq100 at Dhulikhel Hospital Kathmandu University Hospital. This case series revealed the presence of Enterovirus C (coxsackievirus strain A24) as the major pathogen responsible for the outbreak. This case series contributes valuable insights into the genomic diversity of conjunctivitis-associated viruses, highlighting the potential of mNGS in enhancing diagnostic capabilities and guiding public health responses.

## Background

Conjunctivitis is the inflammation of the conjunctival tissue of the eye along with engorgement of the blood vessels, ocular discharge and pain. [1,2] The causes of conjunctivitis have been known to be of viral, allergic and bacterial of origin; and could be infectious or non-infectious. [2] Viral conjunctivitis, also known as Pink Eye or Eye Flue, constitute of 75% of the cases can produces red, irritated eye with watery discharge, with occasional presence of preauricular adenopathy. [3,4] Around 90% of all viral conjunctivitis is caused by adenovirus. [5] Bacterial conjunctivitis, on the other hand, has similar presentation to viral but is less common and has more mucopurulent drainage. [3,6] The bacterial conjunctivitis is mostly caused by *Staphylococcus spp*., *Haemophilus influenza, Streptococcus spp, Moraxella catarrhalis* and other gram-negative bacteria. [7]

Infective conjunctivitis can be transmitted by direct contact, abnormal proliferation of conjunctival flora, contaminated fingers or discharge. [2,8,9] It has been found that children are most susceptible to viral infections while adults tend to get more bacterial infections. [2] In terms of chronicity, conjunctivitis can be divided into acute with rapid onset and duration of four weeks or less, subacute, and chronic with duration longer than four weeks. [10] In severe conjunctivitis, the individuals are extremely symptomatic with abundance of mucopurulent discharge.[5] Some conjunctivitis cases developed keratits and take a longer time to heal and develop long term visual complications like corneal scarring.

The resolution of conjunctivitis can usually take up to 3 weeks. [1,5] In case of viral conjunctivitis, the treatment is aimed at symptomatic relief instead of eradication of the infection with frequent use of preservative-free artificial tears for lubrication, dilatation of allergens and flushing the ocular surface clean from many inflammatory mediators. [2] In bacterial conjunctivitis, topical antibiotic drops used and, in some instances, steroids and non-steroidal anti-inflammatory drugs are given judiciously. [19] Prescribing unnecessary antibiotics for treatment of viral conjunctivitis is one of the major costs of any healthcare system and can add on to the global problem on antibiotic resistance. [2,11] Conjunctivitis can have a considerable economic and societal impact, due to the costs of visits to the ophthalmologists, cost of diagnostic tests, treatment, and time lost from work or school.

Diagnosis of conjunctivitis is usually clinical and laboratory testing is typically not indicated unless the symptoms are not resolving and infection last longer than 4 weeks. [2] [12] In case of clinical manifestation of adenoviral conjunctivitis, cell culture has been proved to be gold standard. [13] Recently, molecular tests have been used for diagnosis with considerable specificity and sensitivity. [13,14] Next generation sequencing approaches, on the other hand, have revealed diverse microbiome profile with some techniques being able of profiling to species level and is inherently unbiased and hypothesis-free. [15-18]

Since July 2023, Nepal had an epidemic outbreak of conjunctivitis cases across the country. The surge was so high that more than 60% of the OPD patients in various eye hospitals of Nepal were filled with conjunctivitis cases and schools were shut down to prevent the community transmission. The cases mainly presented as pink eye and viral origin was the top suspicion.[20] This rapid outbreak mostly affected children and people with low immunity.

## Methods

Himalaya Eye Hospital, Pokhara, Nepal experienced a conjunctivitis outbreak from mid-July until September 2023. The conjunctival swabs were collected from the subjects (2-55 years) visiting HEH, as part of routine diagnosis, at the first day of ocular examination. These swabs were aliquoted in RNA/DNA Shield and transported (n=25) in cold chain to DHKUH. At DHKUH, the swabs were extracted for DNA and RNA (Zymo Quick DNA/RNA Pathogen Miniprep Kit, Cat: R0142) followed by subsequent DNA and RNA library preparation (New England Biolabs) for metagenomic next generation sequencing. The respective libraries were pooled and quality checked by Agilent Tapestation and then loaded in Illumina iSeq100, with 4 million reads at 2x150 bp length. Additional process controls (extraction and library preparation) and sample controls (normal flora) were included for background model. The downstream metagenomic investigation was done by CZID (Illumina mNGS Pipeline v8.2), according to the read scores and Z-score for the genus compared with the non-AES and no-template controls.

## Results

During the outbreak season from July to September 2023, 49,544 subjects visited Himalayan Eye Hospital with conjunctivitis diagnosed (clinically) in 14,926 subjects (30.1%). The detailed case distribution during this period has been presented in Table 1.

**Table 1:**
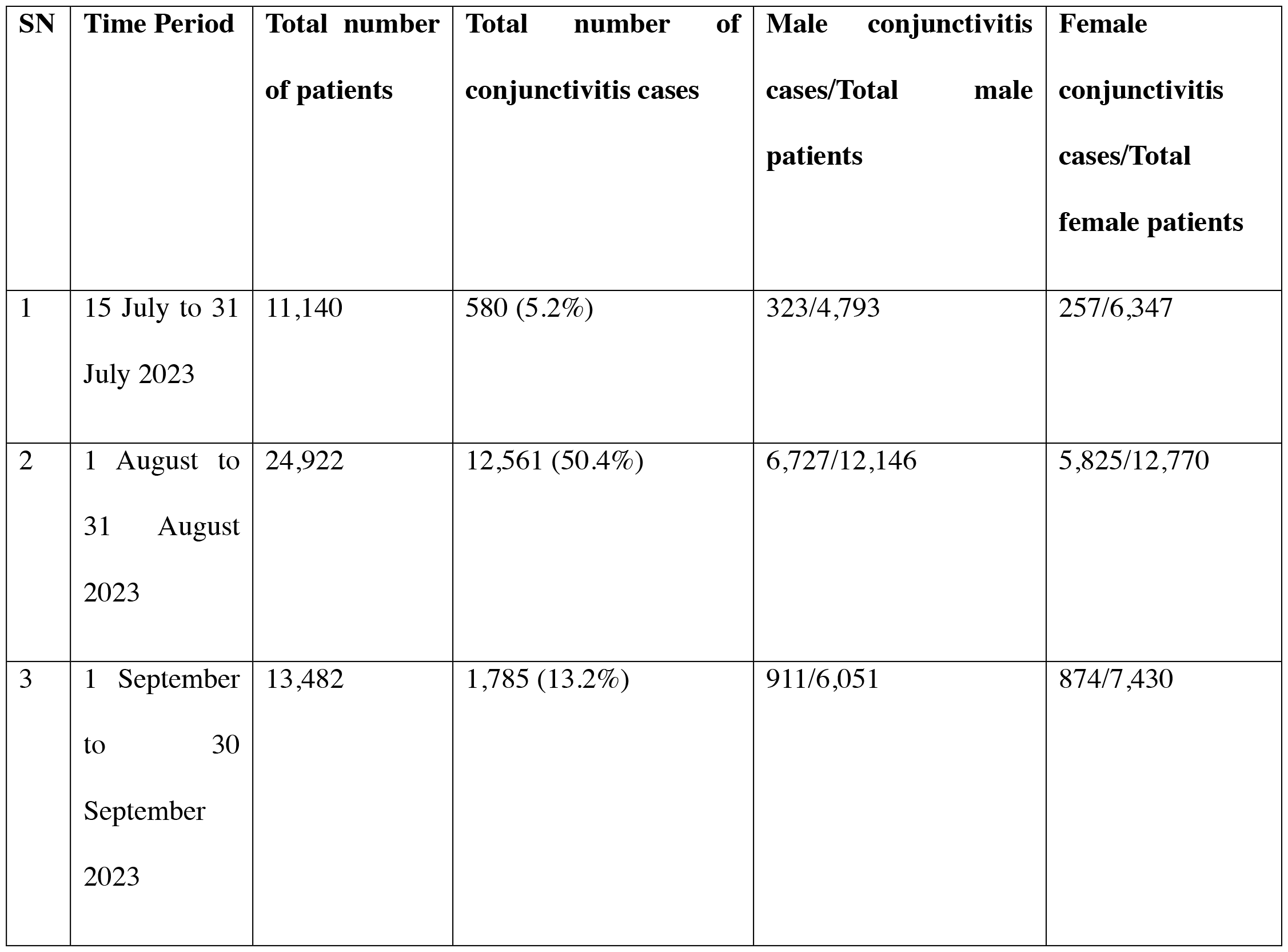
Case distribution during the conjunctivitis outbreak from July-September 2023, in Himalaya Eye Hospital, Pokhara.

The majority of the cases presented with sudden onset of foreign body sensation with redness in one eye followed by involvement of the other eye in 1-2 days. There was presence of watery discharge, minimal pain, occasional lid swelling but prominent redness in some part of conjunctiva suggestive of acute hemorrhagic conjunctivitis. The management were done with the use of ocular lubricants, care of personal hygiene and topical antibiotics as per need.

Among the conjunctival swabs received at DHKUH, samples from 22 subjects were evaluated at DHKUH using unbiased metagenomic next generation sequencing.

As seen in figure 1, most of the samples had *Cutibacterium acnes* and *Escherichia coli*. We have also found this organism as part of healthy conjunctival flora, as per our previous studies (unpublished data). Other organisms observed that could likely cause conjunctivitis, were *Micrococcus luteus* and *Sphingomonas spp*. Enterovirus C (coxsackievirus strain A24) was seen in total of 11 samples (Figure 2), while four samples (S3, S9, S16, S21) had coverage width >95% for A24 strain, as seen in figure 3.

**Figure 1:**
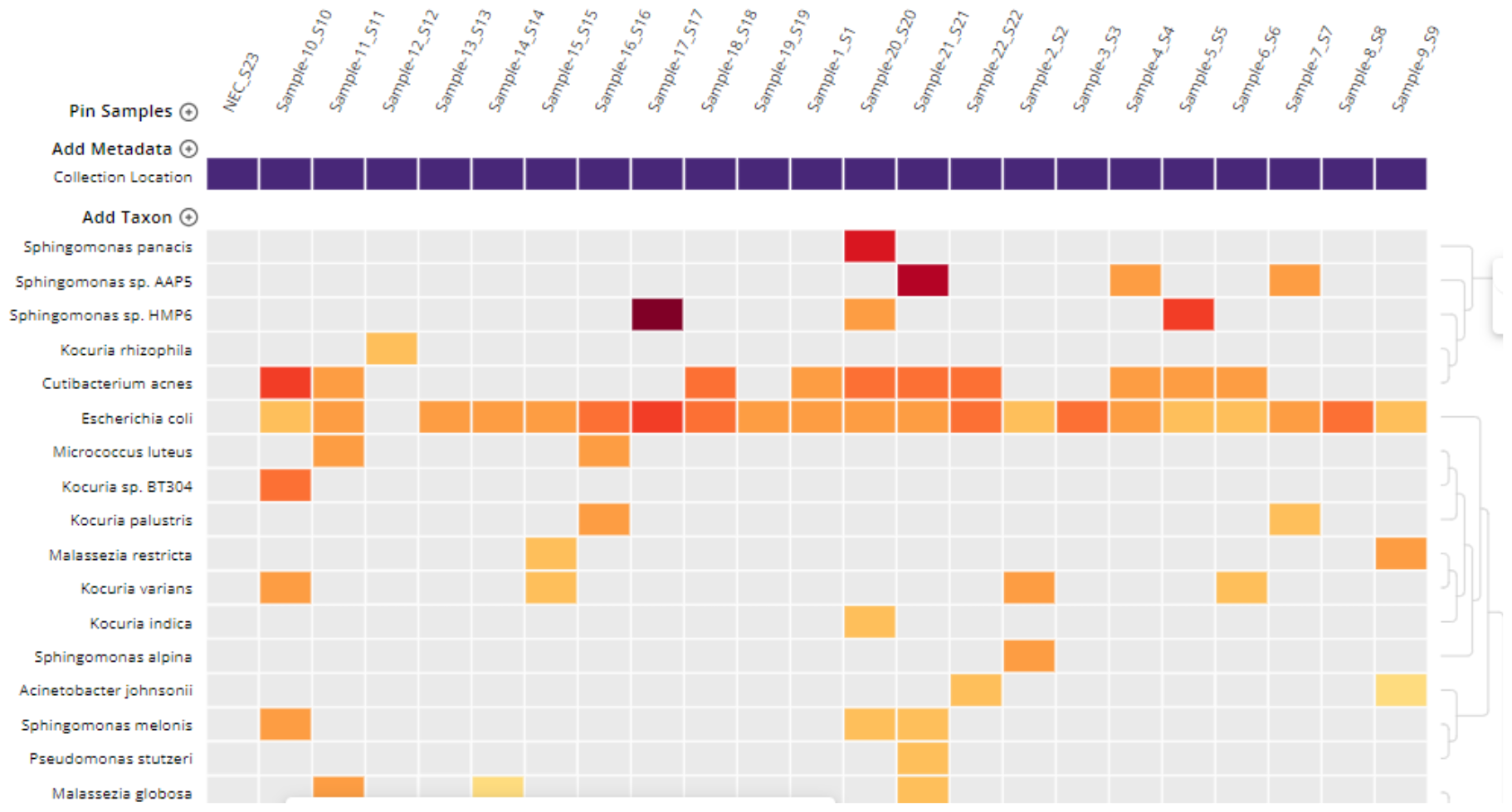
Heatmap of libraries for S1 to S22: The organisms (at the genus level) that were seen in the samples are shown on x axis, while the names of the samples are on the y axis. NEC in the figure stands from Negative Extraction Control, while NLC (Negative Library Control) did not enough reads to be seen in the heatmap. This heatmap used the background model from conjunctival samples of healthy controls with threshold of NT rPM (nucleotide reads per million) >=10 and NT L (alignment length in basepairs: length of the aligned sequence)>=50

**Figure 2:**
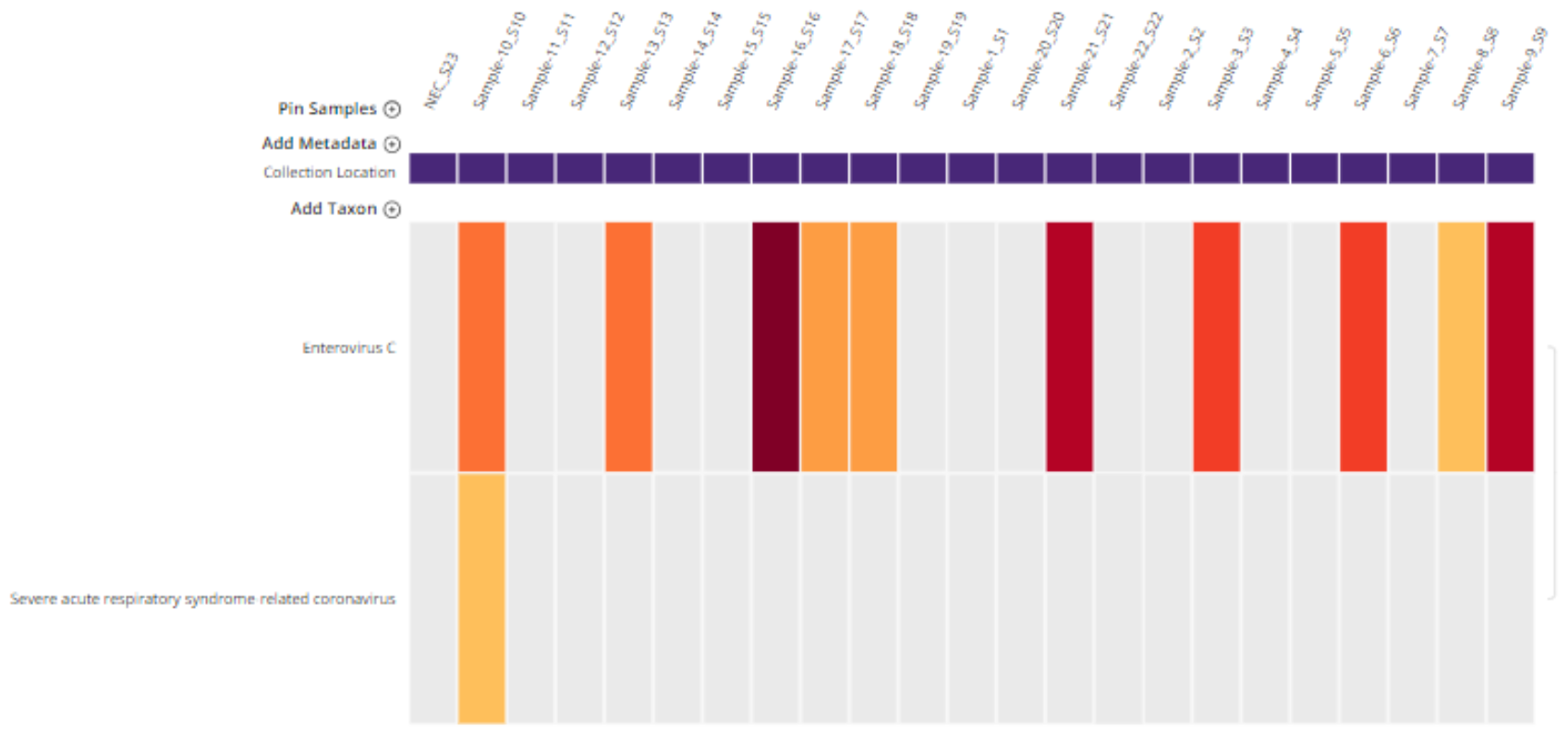
Heatmap of top hits, of viral origin, from sequencing of libraries for S1 to S22. This heatmap used the background model from eye samples of healthy controls with the threshold of NT rPM (nucleotide reads per million) >=10 and NT L (alignment length in basepairs: length of the aligned sequence)>=50.

**Figure 3:**
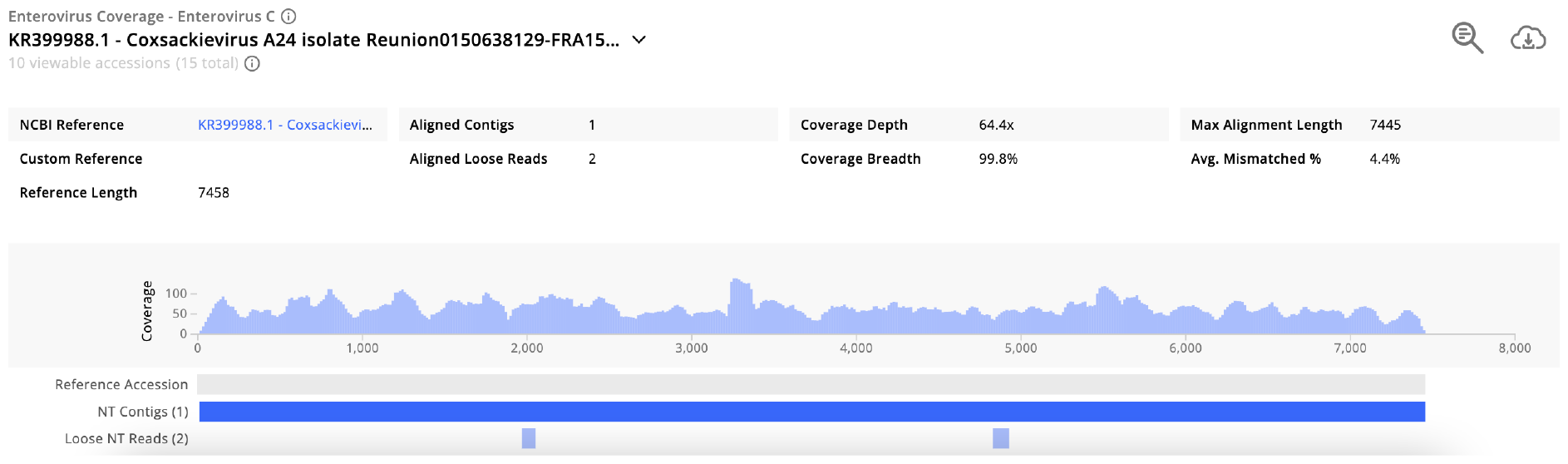
Coverage visualisation of sample S16 for Enterovirus C.

**Figure 4:**
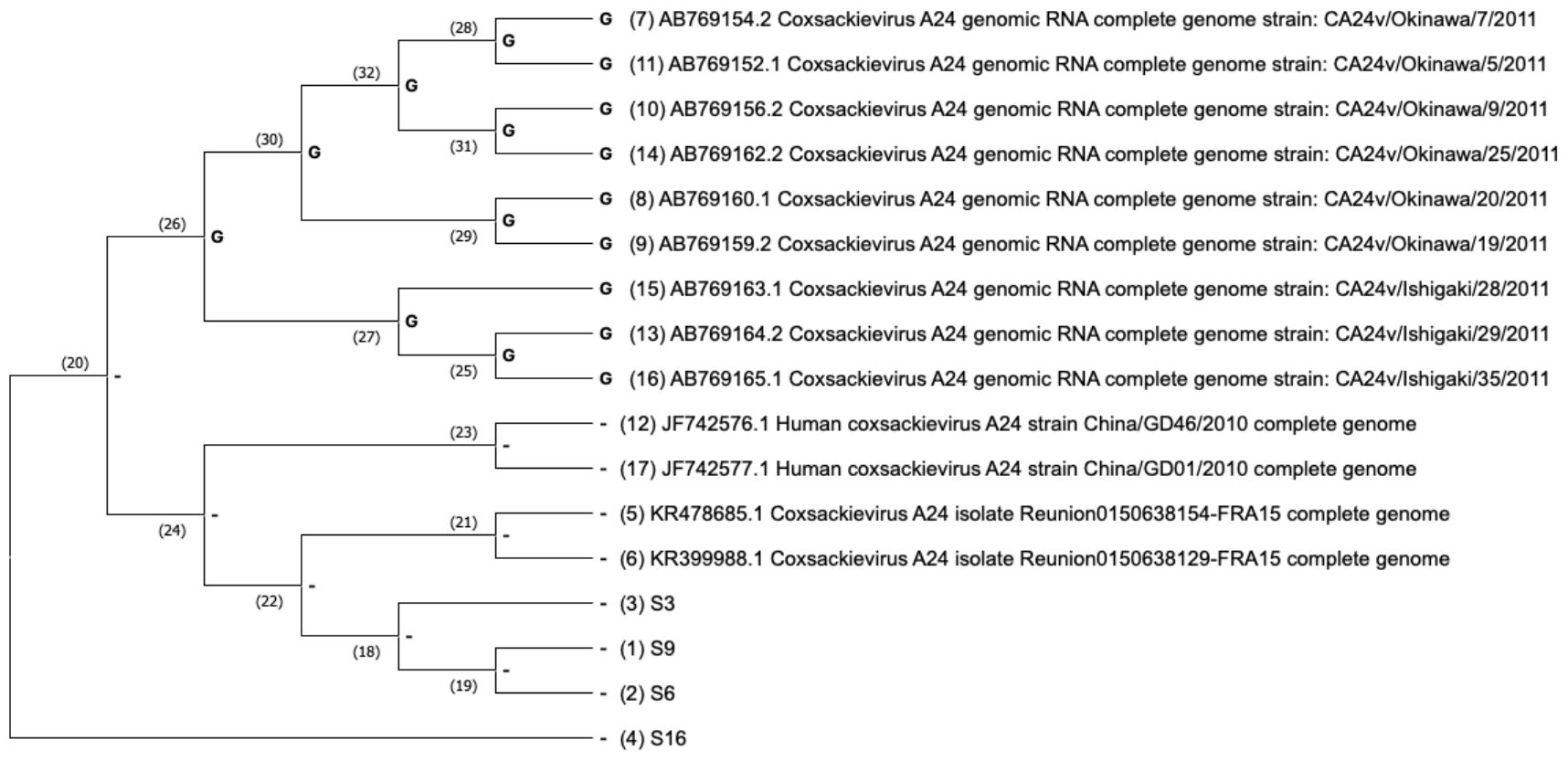
Phylogenetic analysis of four high coverage samples (S3, S6, S9, S16) against other coxsackievirus genomes, from NCBI, reported to have caused ocular infections.

The phylogenetic analysis of the consensus Enterovirus C genomes showed variation from other reported ocular infections associated coxsackievirus (from studies in China, Japan and France). Additionally, samples S3, S6 and S9 slightly had some common genomic regions with coxsackieviruses isolates from France. The phylogeny was made using maximum likelihood statistical method (Mega X), Tamura-Nei model with nearest neighbor interchange as the maximum likelihood heuristic method.

## Discussion

The recent outbreak of conjunctivitis started in mid-July, peaked in August with half of the cases visiting the hospital, suffering from conjunctivitis. [20] This prevalence then decreased in September 2023.

The major pathogen causing the mass outbreak of conjunctivitis in 2023 was found to be *Enterovirus C* (coxsackievirus strain A24). Enterovirus family is one of the most common causes of viral conjunctivitis.[25] However, this particular strain A24, first observed in 1970, is known to cause highly contagious conjunctivitis and has a short incubation period of 12 hours to 3 days. [26, 27] This might be the reason behind sudden spike of cases in Nepal as this same strain has previously been reported to cause several outbreaks throughout the world, with one study revealing import of the virus from Asia. [28, 29]

*Cutibacterium acnes*, which is a normal flora and an opportunistic infectant was seen.[21] The samples also had *E. coli*, which is known to cause eye infections such as conjunctivitis. Two samples (S11 and S16) had hits for *Micrococcus luteus*, known to cause keratitis, which needs to be further confirmed through polymerase chain reaction (PCR). [23] Few samples also had *Sphingomonas spp* in them. Studies have shown that certain *Sphingomonas spp*. cause eye infections.[24] Interestingly, one of the subjects also showed SARSCOV2 in the conjunctival swab. Though some studies have found evidences of SARSCOV2 positive conjunctivitis, this could be remnants of respiratory COVID infection in the subject because this particular hit had only few reads. [31]

Our findings correlated to results of PCR tests from National Public Health Laboratory (NPHL), which found enterovirus in eight of ten samples collected from a hospital in Lalitpur, Nepal.[30] However, the sub type and strain was not identified or reported in their analysis. The unbiased mNGS approach, in our investigation, was able to find numerous possible organisms, of viral or bacterial origin, in a single evaluation, including the strain. The targeted molecular approaches such as PCR, on the other hand, require prior information of genetic material and microbiological methods are only able to identify the presumed cause of ocular infection in about 40% cases. [32] Furthermore, the sequencing approach also visualizes the variability of the microbiome. [32, 33] As seen, we were able to identify the diverse organisms observed and additionally identified the sub-type and strain of Enterovirus C, the coxsackievirus A24. These A24 strains of ENV-C were, in general, genomically different from other reported ocular infections associated coxsackievirus (from studies in China, Japan and France). [34-36] However, samples S3, S6 and S9 slightly had some common genomic regions with coxsackieviruses from a study in France. [34] This particular study also reported these viruses being involved in a major conjunctivitis outbreak in Reunion region of France in 2015, with red eye being the most common symptom, similar to our investigation. [34]

Thus, our investigation depicted the strain of enterovirus (A24) responsible for conjunctivitis outbreak in Nepal and supported the use of unbiased mNGS approach for deeper investigation of ocular infections, for instance in identification of viruses to strain level.

## Data Availability

All data generated or analyzed during this study are included in this article.

## Declarations

### Ethics approval and consent to participate

This study was ethically cleared from IRC-KUSMS. This study directly did not contact the human subjects and investigated stored conjunctival samples and secondary clinical and demographic metadata.

### Consent for publication

Not Applicable.

### Availability of data and materials

All data generated or analyzed during this study are included in this article.

### Competing interests

The authors declare that they have no competing interests.

### Authors’ contributions

Conceptualization: Rajeev Shrestha, Ranju Kharel, Eliya Shrestha; Methodology: Rajeev Shrestha and Nishan Katuwal; Investigation: Nishan Katuwal and Aastha Shrestha; Resources: Rajeev Shrestha, Nishan Katuwal; Data curation: Eliya Shrestha and Nishan Katuwal; Writing-original draft preparation: Eliya Shrestha and Nishan Katuwal; Writing-review and editing: all authors; Supervision: Rajeev Shrestha; Project administration: Rajeev Shrestha. All authors have read and agreed to the published version of the manuscript.

## Acknowledgments

We gratefully acknowledge the participants and research staffs from Himalaya Eye Hospital and Dhulikhel Hospital Kathmandu University Hospital.

## Notes

### Competing Interest Statement

The authors have declared no competing interest.

### Funding Statement

This study did not receive any funding.

